# Overcrowding and Exposure to Secondhand Smoke Increase Risk for COVID-19 Infection Among Latinx Families in Greater San Francisco Bay Area

**DOI:** 10.1101/2021.01.19.21250139

**Authors:** Andrea DeCastro Mendez, Milagro Escobar, Maria Romero, Janet M. Wojcicki

## Abstract

**Background:** The novel coronavirus (COVID-19) has disproportionately impacted the Latinx community in the United States. Environmental risk factors, including community level pollution burden and exposure to smoking and secondhand smoke, have not been evaluated in relation to risk for infection with COVID-19.

**Methods:** We evaluated self-reported infection rates of COVID-19 in three, preexisting, longitudinal, Latinx family cohorts in the San Francisco Bay Area from May through September 2020 (N=383 households, 1,875 people). All households were previously recruited during pregnancy and postpartum at Zuckerberg San Francisco General Hospital (ZSFG) and UCSF Benioff before the pandemic. For the COVID-19 sub-study, participants responded to a 15-minute telephonic interview where we assessed food consumption patterns, housing and employment status, and history of COVID-19 infection based on community and hospital-based testing. We also evaluated secondhand smoke exposure based on previously collected self-reported data. Environmental pollution exposure was determined from census tract residence using California’s EnviroScreen 2.0 data. Non-parametric tests and multiple logistic regression were used to assess possible associations and independent predictors of COVID-19 infection.

**Results:** In the combined Latinx, Eating and Diabetes Cohort (LEAD) and Hispanic, Eating and Nutrition (HEN) cohorts there was a 7.6% household infection rate (14/183) with a lower rate of 3.5% (7/200) in the Telomeres at Birth (TAB) cohort. Larger household size increased risk for infection (OR, 1.43 (95%CI 1.10-1.87)) in the combined LEAD/HEN cohorts and increasing number of children trended towards significance in the TAB cohort (OR 1.82, 95% CI 0.98-3.37). Any exposure to secondhand smoke in the household also trended towards increasing risk after adjusting for household size and other exposures (OR 3.20, 95%CI 0.80-12.73) and (OR 4.37, 95% CI 0.80-23.70). We did not find any associations between neighborhood pollution level based on census track location and risk of infection. Furthermore, we found weak evidence between dietary exposure and risk of COVID-19 infection after adjusting for possible confounders.

**Conclusion:** Crowding as indicated by household size increases risk for COVID-19 infection in Latinx families. Exposure to secondhand smoke may also increase risk for COVID-19 through increased coughing, respiratory impairment and increased travel of virus on smoke particles. Public policy and health interventions need to ensure that multiunit residential complexes prevent any exposure to secondhand smoke.

## Background

### COVID-19 in USA Latinx

The novel Coronavirus (COVID-19) has infected over 23.6 million individuals in the United States and more than 394,495 have died (Centers for Disease Control and Prevention, 2020). COVID-19 disease disproportionately affects US racial and ethnic minorities (Tai et al., 2020). The Latinx community comprises 18% of the United States population but 33% of the confirmed COVID-19 cases and 20% of all deaths (Rodriguez-Diaz et. al., 2020). Pediatric COVID-19 infection rates are also higher in US racial and ethnic minorities (Ranabothu et al., 2020). As of fall 2020, there were 161,387 school-aged children with COVID-19 in the United States, 42% of whom were Latinx children compared to 32% non-Hispanic White and 17% non-Hispanic Black (Leeb et al. 2020). In California, Latinx’s children account for 47.9% of the state’s children, yet represent 68.5% of all child COVID-19 cases and 50% of deaths (California Department of Public Health, 2020).

### Risk Factors in Latinx communities

Latinx adults comprise a large proportion of the country’s workforce but hold mostly low-paying employment lacking benefits (Tran, 2019). When COVID-19 shelter-in-place began, 16.2% of Latinx workers had the option to work from home in comparison to the 29.9% of White workers (US Bureau of Labor Statistics, 2019). Household vulnerability increases risk for COVID-19 infection based on factors, such as crowding and the presence of multi-generational households (Khazanchi et al., 2020). California Latinx households are more likely to reside in homes with non-family members compared with non-Latinx households (Latino Caucus, 2017).

### COVID-19 Environmental and Dietary Factors

Extended exposure to air pollution, including secondhand smoke, can increase infection risk and mortality from COVID-19 by compromising respiratory health and increase risk for chronic cough (Costello et. al., 2020; Fell et al., 2018; Roth, 2020; Harvard TH Chan School of Public Health, 2020). Communities of color, including Latinx families, are more likely to be residing in areas of high pollution than White counterparts (American Lung Association, 2020).

Shelter-in-place orders have also adversely impacted global nutritional health as individuals are more likely to be consuming poorer-quality diets with more unhealthy eating practices (Ghosh, 2020; Khubchandani et al., 2020). Poor nutrition, including low levels of Vitamin D and selenium, can increase susceptibility to illness including potentially risk of novel coronavirus infection (Rhodes et. al, 2020; Bae et al., 2020). Western nutrition consisting of foods high in saturated fats, sugars and refined carbohydrates activate the innate immune system while deactivating the adaptive immune system resulting in inflammation and diminished defense against viruses, including potentially COVID-19 (Butler et. al, 2020).

In our COVID-19 risk study, we sought to assess the relationship between environmental risk factors including secondhand smoke exposure and nutritional health and COVID-19 infection in an urban, inner-city Latinx population.

## Methods

### Recruitment

Families in the present study are part of three, longitudinal cohort studies of Latinx families: the Hispanic Eating and Nutrition Study (HEN) (original N=201) (Kjaer et al, 2019; Wojcicki et al., 2011), Latinx, Eating, and Diabetes Study (LEAD) (original N=97) (Ville et al., 2017; Wojcicki et al. 2018), and the Telomeres at Birth Study (TAB) (original N= 424). HEN and LEAD mothers were recruited during pregnancy primarily at Zuckerberg San Francisco General Hospital (ZSFG) with a minority from UCSF Benioff whereas TAB participants were recruited in the post-partum unit before hospital discharge (mostly at UCSF Benioff and the remaining at ZSFG). Participants for these three studies have been followed over the course of 13, 8, and 1-2 years, respectively. Specifics of the studies have previously been described (Kjaer et al., 2019; Wojcicki et al., 2011; Ville et al., 2017; Wojcicki et al., 2018).

We contacted participants in HEN, LEAD, and TAB telephonically from May through September 2020 to partake in this COVID-19 sub-study. Participants gave their language preference (Spanish or English) and were told information about the COVID-19 study by the bilingual research coordinators (ADM, ME and MR) and the principal investigator (JW). Those who chose to participate gave verbal consent for the 15-minute interview. We assessed COVID-19 symptoms and lab-based testing, food consumption patterns during COVID-19, and current employment and housing status. Participants received a **$**15 gift card to Amazon or Target as compensation for study participation. The Committee on Human Research (CHR), the UCSF Institutional Review Board approved all aspects of the study.

We used the CalEnviroScreen 3.0 (California Office of Environmental Health Hazard Assessment, 2018) to assess levels of pollution by census tract in the state of California. Specifically, CalEnviroScreen 3.0 determines California communities disproportionately affected by pollution by examining pollution burden score and percentile.

### Procedures

All participants were asked if they had symptoms consistent with COVID-19 from the start of lockdowns in March 2020 until the time of the interview (May-September 2020). COVID-19 symptoms that we asked about included cough, fever, muscle aches, stomach-ache, diarrhea, vomiting, sore throat, shortness of breath, COVID toes or loss of smell or taste.

All participants were asked if anyone in their household had undergone a COVID-19 test at a doctor’s office, hospital or testing site. Households were determined to be positive for COVID-19 if anyone reported a positive polymerase chain reaction (PCR test), and/or symptoms including loss of taste/smell. Other questions focused on household crowding including household size, number of bedrooms and bathrooms and number of household members sharing bedrooms and bathrooms. We also assessed the number of working days each week for household members, and COVID-19 specific risk factors including the use of face masks in public and the frequency of public transportation use.

At the baseline assessment of all families, we collected maternal primary language use (English, Spanish, or bilingual) and education level (primary or less than a high school diploma, high school diploma, some college or more). We also annually collected information on exposure to secondhand smoke in the household. We were able to use the address of residence at the time of the COVID-19 sub-study to find the CalEnviroScreen3.0 score for each participant to identify the extent to participants were impacted by pollution including PM2.5 exposure, pesticide use, diesel particulate matter and drinking water contamination (California Office of Environmental Health Hazard Assessment, 2018).

### Statistical Analysis

We combined HEN and LEAD cohorts for a total of 183 participants for analysis as recruitment specifics and demographics of these cohorts are similar (both recruited at ZSFG primarily with similar exclusion and inclusion criteria) (Kjaer et al., 2019; Wojcicki et al., 2011; Ville et al., 2017; Wojcicki et al., 2018). The TAB cohort was analyzed separately and only participants who identified as Latinx were included in the analysis (n=200). Data was assessed for normality using graphical tests of normality and statistical tests including the Shapiro-Wilk and the Kolmogorov-Smirnov tests. As the data were not normally distributed, we used non-parametric tests of association including the Wilcoxon rank-sum test and the chi-square test to assess measures of association for continuous and categorical data.

Variables that were significant in bivariate analysis at p<0.10 were included in multivariable analyses. Other variables that have been shown to be associated with COVID-19 infection based on biological plausibility were also included in models even if they did not meet the statistical threshold. We ran separate analyses for the HEN/LEAD versus the TAB cohorts. We conducted secondary multivariable analyses for both HEN/LEAD and TAB that included dietary factors that were significant at p<0.20 as one of our primary outcomes of interest was the relationship between nutritional health and COVID-19 infection. These models tended to be smaller as some of the data was missing for food frequency and dietary recall. All analyses were conducted using Stata 15.1.

## Results

Of the 183 families in the combined HEN/LEAD cohorts, there were 14 families with cases (7.6%) of COVID-19 reported. The TAB Cohort had 200 families and 7 cases of COVID-19 (3.5%) (Tables 1 and 4).

### Socio-demographics

Larger household size was associated with COVID-19 infection in HEN/LEAD cohorts 6.2±2.7 versus 4.9±1.6 (p=0.02; Table 1). More family members sharing bedrooms was also associated with increased risk for infection 2.9±1.5 versus 2.5 ± 1.2 (p=0.03; Table 1). Increasing number of people eating together approached statistical significance for COVID-19 infection (p=0.07; Table 1). For the TAB cohort, a higher number of children in the household was associated with increased risk for infection 3.3±1.3 versus 2.4± 1.1 (p=0.03; Table 4). A greater number of individuals eating together was protective against infection (2.7±0.8 versus 3.8±1.2; p=0.01; Table 4). We did not find any association between COVID-19 infection and maternal high school diploma, primarily Spanish language use or Mexican origin (versus Central American or other Hispanic ethnicity) for HEN/LEAD nor the TAB cohorts.

**Table 1:**
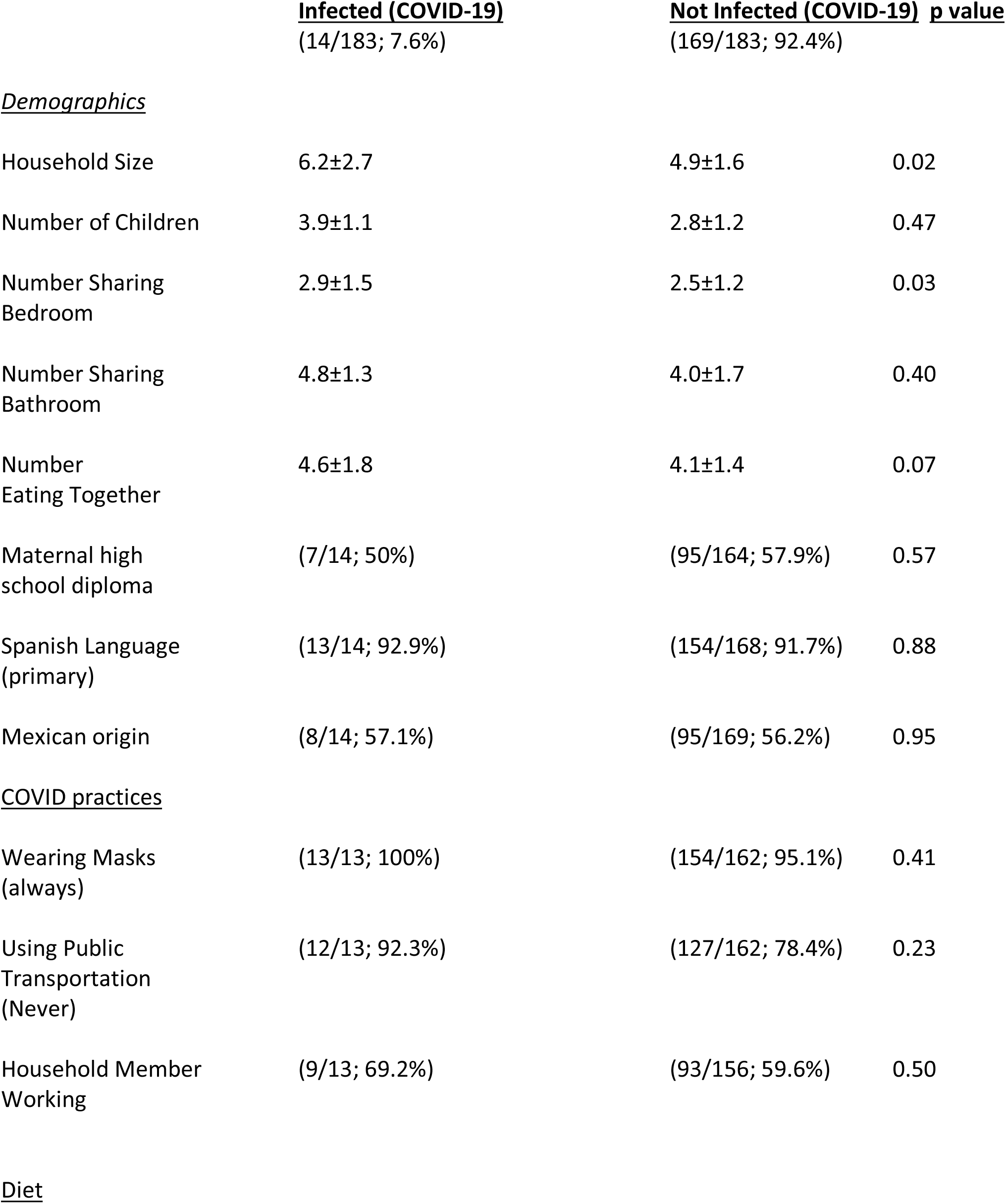
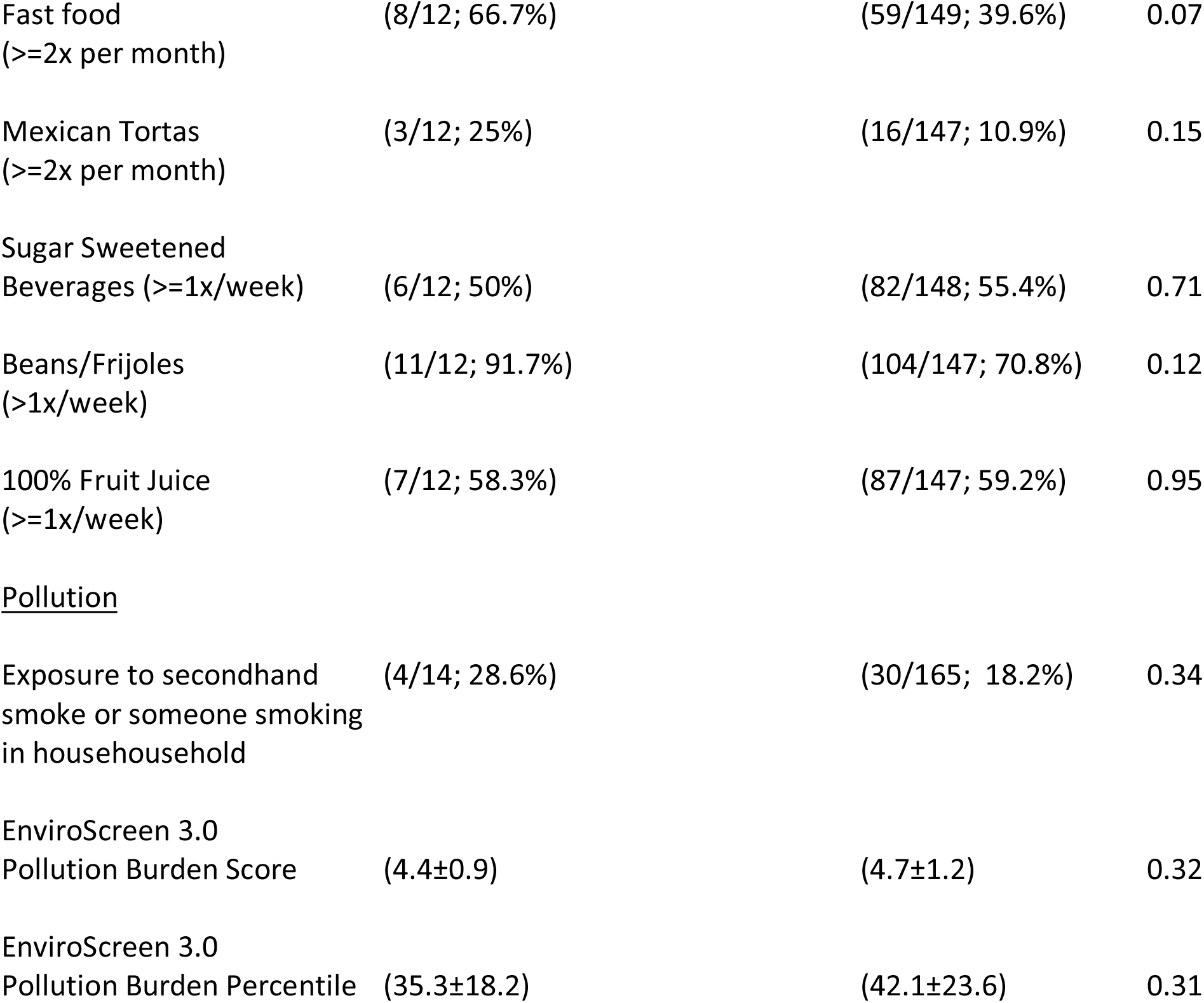
**Family Demographics, Diet and COVID-19 Infection (HEN and LEAD Cohorts Combined)**

**Table 2:** **Independent Household Predictors for COVID-19 Infection: Multivariable Logistic Regression (n=169)**

|  | Odds Ratio (95% Confidence Interval), p-value |  |
| --- | --- | --- |
| Household number | 1.43 (1.06-1.93) | 0.02 |
| Number sharing bedroom | 1.03 (0.66-1.60) | 0.90 |
| Number eating together | 1.10 (0.79-1.55) | 0.57 |
| Any exposure to secondhand smoke | 3.20 (0.80-12.73) | 0.099 |
| Public transportation use (any) | 0.38 (0.05-3.23) | 0.38 |

### COVID-19 Prevention and Risk Associated Practices

We did not find statistically significant differences between COVID-19 prevention practices, such as wearing masks and risk for infection (Tables 1 and 4). There was also no association between COVID-19 and frequency of practices associated with elevated risk such as public transportation use. Having household members working outside of the house trended towards statistical significance in the TAB cohort with a higher percentage of families with a household member infected working outside the home (100% versus 75.1%; p=0.13; Table 4).

### Diet

We found a higher percentage of fast food consumption (>=2x/month) in HEN/LEAD among those infected with COVID-19 with a trend towards statistical significance (66.7% versus 39.6%; p=0.07; Table 1). Higher consumption of Mexican tortas (a fast food sandwich) was also greater among those who were infected versus uninfected (>=2x/month), 25.0% versus 10.9%; (p=0.15; Table 1). Beans were consumed more frequency by families with COVID-19 infection compared to those families without COVID-19 infection (91.7% versus 70.8%; p=0.12; Table 1).

In the TAB cohort, there was a trend towards higher consumption of 100% fruit juice among those infected with COVID-19 versus uninfected although results were not statistically significant (66.7% versus 36.8%; p=0.14; Table 4). Other dietary intake factors including consumption of fast food, sugar sweetened beverages and 100% fruit juice were not associated with COVID-19 infection.

### Secondhand Smoke Exposure and Environmental Pollution

There was a higher percentage of exposure to secondhand smoke in HEN/LEAD in those families with COVID-19 infection (28.6% versus 18.2% for those uninfected) although the differences were not statistically significant in bivariate analysis (p= 0.34; Table 1). We did not find any association between pollution burden score or pollution burden percentile as indicated on the Enviroscreen 3.0 related to COVID-19 infection (Table 1)

In TAB, a higher percentage of those with exposure to secondhand smoke had someone in the household with COVID-19 (42.9% versus 11.4%; p=0.01; Table 4). There was no association between pollution burden score or percentile as per Enviroscreen 3.0 and COVID-19 infection (Table 4).

### Multivariable findings

Independent predictors for COVID infection in HEN/LEAD include a higher number of household members in the house (OR 1.43,95% CI 1.06-1.93; p= 0.02; Table 2). Increased exposure to secondhand smoke trended towards statistical significance after adjusting for other factors including number of household members, number of household members eating together and number sharing a bedroom (OR 3.20, 95% CI, 0.80-12.73; p= 0.099; Table 2).

For the TAB cohort independent predictors for COVID-19 infection included a higher number of household children (OR 1.82, 95% CI 0.98-3.37; p=0.06). Exposure to any secondhand smoke trended towards statistical significance (OR 4.37, 95%CI 0.80-23.70; p=0.09; Table 5). There was no association between risk of infection and any use of public transportation and maternal education after adjusting for exposure to secondhand smoke and number of the children in the household.

We did not find any association between dietary intake including consumption of sugar-sweetened beverages, 100% fruit juice, Mexican tortas and beans with risk for COVID-19 infection after controlling for other risk factors including crowding associated ones and secondhand smoke exposure (Tables 3 and 6).

**Table 3:** Dietary Predictors for COVID-19 Infection: Multivariable Logistic Regression (n=154)

|  | Odds Ratio (95% Confidence Interval), p-value |  |
| --- | --- | --- |
| Household number | 1.43 (1.10-1.87) | <0.01 |
| Any exposure to secondhand smoke | 3.07 (0.62-15.08) | 0.17 |
| High tortas consumption ($\geq 2x/mo$) | 2.38 (0.59-9.60) | 0.22 |
| High fastfood consumption ($\geq 2x/mo$) | 4.07 (0.53-31.24) | 0.18 |
| High beans/frijoles consumption ($>1x/week$) | 3.13 (0.54-18.17) | 0.20 |

**Table 4:**
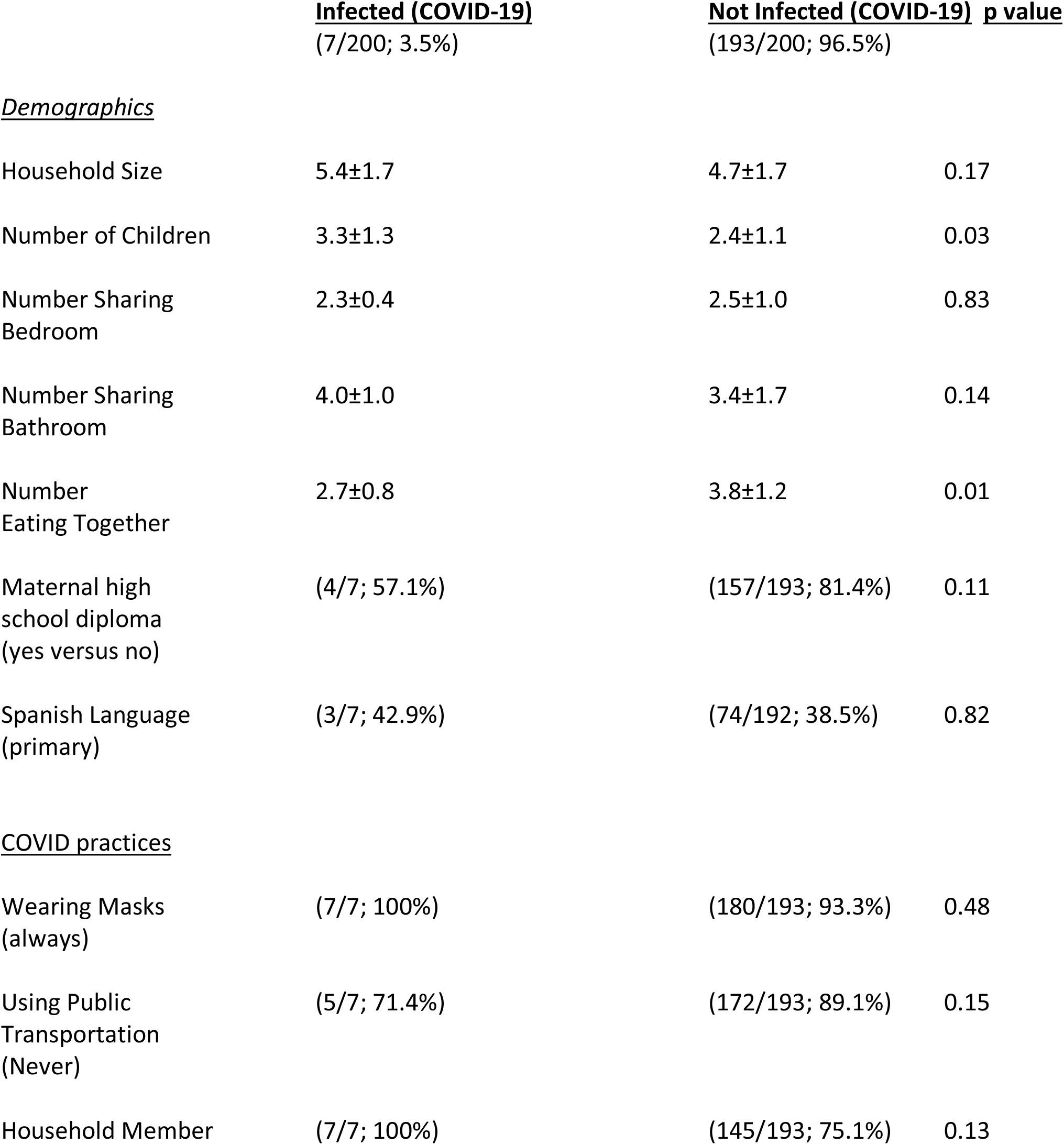
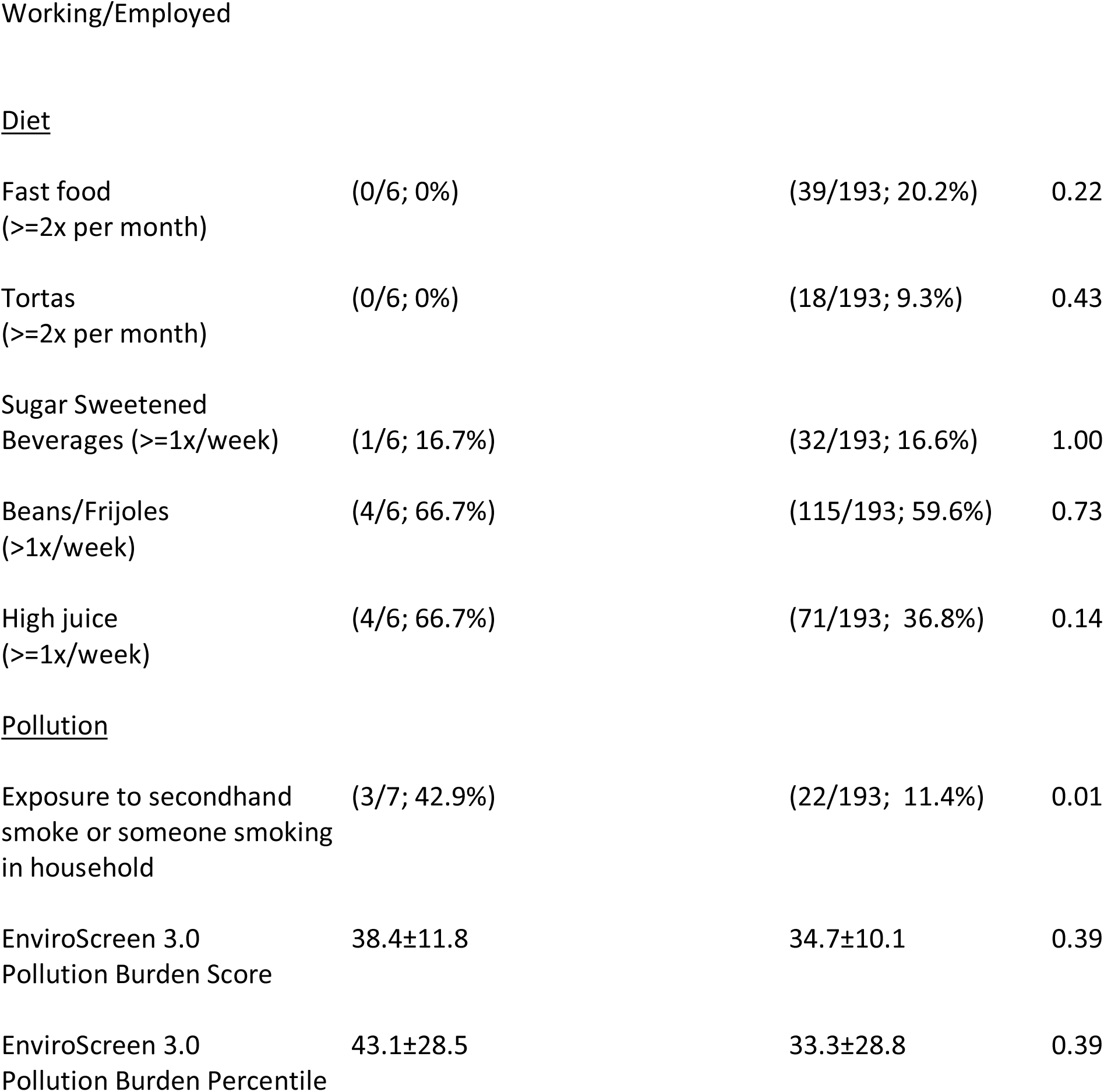
**Family Demographics, Diet and COVID-19 Infection (TAB Cohort)**

**Table 5:** **Independent Household Predictors for COVID-19 Infection: Multivariable Logistic Regression (n=200)**

|  | Odds Ratio (95% Confidence Interval), p-value |  |
| --- | --- | --- |
| Number of children in house | 1.82 (0.98-3.37) | 0.06 |
| Mother has high school diploma | 0.45 (0.08-1.60) | 0.35 |
| Any exposure to secondhand smoke | 4.37 (0.80-23.70) | 0.09 |
| Public transportation use (any) | 2.36 (0.32-17.26) | 0.40 |

**Table 6:** Dietary Predictors for COVID-19 Infection: Multivariable Logistic Regression (n=199)

|  | Odds Ratio (95% Confidence Interval), p-value |  |
| --- | --- | --- |
| Number of children in house | 1.58 (0.86-2.92) | 0.14 |
| Any exposure to secondhand smoke | 4.06 (0.67-24.70) | 0.13 |
| High juice consumption | 3.12 (0.54-18.11) | 0.21 |

## Discussion

### Household Crowding

Similar to previous studies, we found that household crowding is associated with COVID-19 infection. Since the start of the pandemic, most COVID-19 transmission occurs in the context of close interpersonal interactions in the home (Rader et. al, 2020). In counties where there are high levels of overcrowding, there is a strong association between COVID-19 infection and overcrowding in the home (Richwine et. al, 2020). Overcrowding occurs when people in a home surpass the home’s capacity to supply privacy, shelter, and space as per the WHO definition (WHO Housing and Health Guidelines. Geneva: World Health Organization; 2018). Implications of overcrowding include increased likelihood of infection and a greater prevalence of respiratory ailments (California Department of Public Health, 2021). The most vulnerable populations for crowding and housing difficulties include with mental illness, chronic health conditions, history of trauma, immigrants, and ethnic/racial minorities (Anderson et. al, 2013; Braubach, 2011; Dohler et. al, 2016; Jones, 2010). Similar to US-based studies, black, Asian and ethnic minorities in the United Kingdom report increased risk for COVID-19 associated with household crowding (Soltan et. al, 2020). Investment in adequate housing in the San Francisco Bay Area could reduce risk for infection in high-risk populations including parts of the Latinx community, that have a higher rate of household crowding.

### Public Health Measures and Interventions to Reduce Household Crowding

Public health measures to reduce overcrowding have traditionally been at the individual level (Shaw, 2004). Interventions for COVID-19 should minimize the number of contacts people with COVID-19 infection have, particularly household contacts. In China, the government provides COVID-19 infected individuals with space to quarantine outside the home and access to free meals, internet, and telephone, thus limiting COVID-19 household transmission (Lai et. al. 2020). In California, hotels have been used as temporary housing locations for high-risk populations, including homeless and health care workers at risk of COVID-19 infection but these interventions have not extended to the general population (State of California, 2021).

Enforcing housing guidelines and codes may be a mechanism to address overcrowding and ensure appropriate ventilation in units; however, there must be a safety net to help families should they need to be relocated due to housing code violations (Krieger et. al, 2002).

### Secondhand Smoking (SHS) and Environmental Pollutants

Secondhand smoke and environment exposures such as pollution increase risk for respiratory illness (San Francisco Tobacco Free Project, 2016; Zeng et. al, 2019). Secondhand smoke consists of more than 7,000 different chemicals from smoke exhaled by a smoker and additional smoke from the cigarette (Center for Disease Control, 2020; San Francisco Tobacco Free Project, 2016). In the United States, approximately 40,000 deaths per year are attributed to secondhand smoke in non-smokers (San Francisco Tobacco Free Project, 2016). We found increased risk associated with exposure to secondhand smoke in both cohorts although these results only trended towards statistical significance. Future studies are needed with larger sample sizes to confirm our findings. There was no association between COVID-19 infection and environmental pollution exposures as indicated by census track data.

Secondhand smoke may facilitate transmission of COVID-19 as the virus attaches to particles expelled by smokers and can travel longer distances (Curley, 2020). Viral particles can travel up to six times farther than if the virus was simply in the air in the absence of smoke (Curley, 2020). Furthermore, smokers tend to exhale more forcefully, and COVID-19 infected smokers could have more particles pushed from their lungs to travel greater distances (Ries, 2020). Previous studies suggest that Latinx households residing in multi-unit housing are exposed to secondhand smoke at very high levels (Baezconde-Garbanati et. al, 2011) with children being the most vulnerable (Zeng et. al, 2019). With pandemic stay-at-home orders in the greater Bay Area, families may have limited options in a confined space particularly if a household member smokes or there is smoking within a multi-unit residence.

### Public Health Measures to Address Secondhand Smoke

Families living in densely packed urban areas or in multi-housing units that do not restrict smoking can be impacted by secondhand smoke exposure (Pickett et. al, 2006). In California, more than 60 cities and counties have 100% smoke free multi-housing units where smoking is prohibited in privately-owned as well as publicly-owned residences. Bay Area municipalities with 100% smoking free laws include Berkeley, Daly City, South San Francisco, Pacifica, and Palo Alto (African American Nonsmokers Foundation, 2020). About two dozen municipalities in California have laws partially restricting smoking in multi-housing units. Municipalities with partial smoking restrictions are less stringent than the 100% smoking free laws as smoking is restricted in some private units of multi-unit housing, yet do not mandate multi-unit homes to be 100% smoke-free (African American Nonsmokers Foundation, 2020). Municipalities in the Bay Area with partial smoking laws include Fremont, Marin County and Pinole (African American Nonsmokers Foundation, 2020).

Secondhand smoking is a concern in residential housing in San Francisco, where many of our participants live (San Francisco Tobacco Free Project, 2016). In 2010, Health Code Article 19F passed in San Francisco preventing smoking in shared areas in multi-unit buildings but allows smoking in individual units and outdoors on private patios or balconies and on street curbs near multi-unit complexes (San Francisco Tobacco Free Project, 2016). As such secondhand smoke can easily seep between units through walls, ventilation systems, windows and doors. With the growing usage of e-cigarettes, San Francisco has begun prohibiting vaping in any location where there is no usage of traditional cigarettes with the law, SF Health Code Article 19N (2014) as second hand vapor also has respiratory health impacts (San Francisco Tobacco Free Tobacco Laws, 2019). However, second hand vapor can similarly travel between units in multi-unit residential complexes.

In December 2020, San Francisco chose not to revise Health Code Article 19F and as such there is no legislation in San Francisco to prohibit smoking inside private dwellings in complexes with three or more units (Moench et. al, 2020; San Francisco Board of Supervisors, 2020). Although Senate Bill-322 will continue to allow landlords to place smoking bans, residents in many multi-use units will continue to face exposure to secondhand smoke and potential increased risk for COVID-19 in the context of COVID-19 exposure (Moench et. al, 2020; San Francisco Board of Supervisors, 2020). Similarly, other East Bay and Peninsula cities where our participants reside including Oakland, San Jose and Vallejo do not have any legislation restricting smoking in multi-housing units.

In the context of COVID-19 shelter-in-place mandates, families spend an increased amount of time at home, with a greater risk for second-hand smoke exposure in the household if neighbors or other household members smoke. There is greater urgency to reform smoking and housing laws to protect residents in multi-unit dwellings.

### Dietary Risk Factors for COVID-19

We did not find any statistically significant dietary risk factors for COVID-19 after adjusting for crowding indices and exposure to secondhand smoke in multivariable models. In bivariate analyses, increased consumption of beans/frijoles, Mexican tortas, fast food and 100% fruit juice were associated with increased risk for infection. Families consuming more beans/frijoles trended towards having COVID-19 infection more frequently than those eating beans with less frequency possible as a marker of socioeconomic status. Beans are more likely to be incorporated in the diet of individuals who are less socioeconomically privileged than in the diets of people with greater resources (Medina et. al, 2020). Having beans in the home tends to provide a sense of food security (Khor et. al, 2018). Future studies should investigate possible links between frijoles/bean consumption, food insecurity, and COVID-19 infection.

Obesity is a major risk factor for COVID-19 and has shown to increase the risk of infection and overall death as increased adiposity negatively impacts immune function and host defenses (Popkin et. al., 2020; Milner et al., 2012). Latinx adults have higher rates of obesity compared with whites; 44.8% versus 42.2% (CDC, 2020). As the novel coronavirus pandemic resulted in changing eating behaviors and decreasing quality of diets in part due to food insecurity, there may be greater risk for infection in families with higher consumption of fast foods, sugar sweetened beverages or other food stuffs associated with obesity and metabolic disease (Khubchandani et. al, 2020). Increased blood sugar levels in non-diabetics has also been linked to more severe symptoms and poorer outcomes of COVID-19, such as ICU admission, ventilation and death (Isler, 2020).

We found no statistically significant relationship between fast food, Mexican tortas and 100% fruit juice consumption and COVID-19 infection, but the families in our cohorts who ate fast food more than twice a month had more COVID-19 infection relative to individuals who ate fast food less frequently. Fast food and 100% fruit juice consumption are associated with increased risk for overweight and obesity and metabolic disease due to the high amount of sugar and fats in these foods and future studies should investigate possible associations with risk for COVID-19 infection and COVID-19 progression (Janssen et. al, 2018). Our sample size was small for this study and future studies should be conducted with larger population groups.

In previous studies, community exposure activities, including restaurant or coffee shop dining, increase risk for COVID-19 infection (Fisher et al, 2020). In a study using cellphone data from 10 urban areas, restaurants were one of the riskiest places for COVID-19 transmission (Carey, 2020). Adults testing positive for COVID-19 infection were twice as likely to have dined at a restaurant 14 days before coming ill compared to those testing negative (Fisher et. al, 2020). Consumption of fast food, such as Mexican tortas or American style fast food, may be synonymous with increased exposure to individuals outside the household or may suggest risk associated with eating at restaurants.

## Limitations and Conclusions

Limitations of this study include small sample size and number of COVID-19 infections. We also did not directly test participants for COVD-19 but relied on self-reported results including recall of COVID-19 related symptoms. Our assessment of secondhand smoke exposure was collected annually and not directly before the start of COVID-19 in some cases and environmental pollutant exposure was based on census track location, not at the household or individual level. Lastly, we did not directly assess other forms of tobacco exposure including the effects of e-cigarettes (vaping) on COVID-19 infections. While the prevalence of cigarette smoking including vaping is lower among Latinxs than other racial and ethnic groups, the odds of smoking increases with acculturation among the Latinx community (Centers for Disease Control and Prevention, 2019; Wang et. al, 2019). Future COVID-19 policy measures in the San Francisco Bay Area should include better implementation of housing guidelines to assure adequate space and ventilation as well as greater inclusion of all individuals for benefits related to healthy living, like hotel housing following COVID-19 infection. Additionally, policy on secondhand smoke exposure should restrict any exposures in multi-unit dwellings by banning any smoking in multi-unit dwellings in San Francisco and other Bay Area cities that do not have existing smoking bans.

## Data Availability

Data are available on request

## Acknowledgements

This project was supported by the generosity of Eric and Wendy Schmidt by recommendation of the Schmidt Futures program, through Covid Catalyst at the Center of Emerging and Neglected Diseases.

